# Estimating Sensitivity from Tandem Colonoscopy Meta-analyses

**DOI:** 10.64898/2026.09.25.26364039

**Authors:** Pedro Nascimento de Lima, Rachel B. Issaka, Carolyn M. Rutter

## Abstract

**Background:** Tandem colonoscopy meta-analyses estimate adenoma miss rates, which have been used as a proxy for colonoscopy sensitivity. Doing so overstates sensitivity because it ignores lesions missed by both exams.

**Methods:** We introduce Bayesian and frequentist approaches to estimate white-light colonoscopy (WLC) sensitivity from tandem colonoscopy meta-analyses. Neither approach assumes that the second colonoscopy is perfect. We apply both approaches to reanalyze data from a recent tandem colonoscopy meta-analysis, overall and across ten subgroups. Sensitivity analyses assess how assumptions about the relative performance of enhanced colonoscopy versus WLC affect estimates.

**Results:** Under our Baseline specification, overall WLC sensitivity estimated by the Bayesian model was 0.54 [0.41, 0.63] (posterior mean and 95% credible interval). Sensitivity rose with lesion size: 0.49 [0.35, 0.60] for 1–5 mm adenomas, 0.64 [0.43, 0.78] for 6–9 mm adenomas and 0.84 [0.70, 0.93] for lesions 10 mm or larger. The frequentist plausible range encompassed the Bayesian estimate. The model-implied miss rate (0.35 [0.30, 0.40]) matched the original meta-analysis estimate and was stable across prior specifications. Assuming enhanced colonoscopy was perfect overstated overall sensitivity by 0.12 [0.06, 0.20].

**Limitations:** Tandem colonoscopy data identify the miss rate but not WLC sensitivity, so estimates depend on an external assumption about the relative performance of enhanced colonoscopy versus WLC. As a result, WLC sensitivity estimates varied with the prior specification, especially for small and flat adenomas.

**Conclusions:** WLC sensitivity may be meaningfully lower than previously assumed in CRC screening cost-effectiveness analyses. Modelers should use colonoscopy sensitivity estimates that do not assume a second colonoscopy exam is perfect.

## 1 Introduction

The effectiveness of colorectal cancer (CRC) screening depends on the sensitivity of colonoscopy, which is used both for primary screening and after an abnormal non-invasive screening test.[1] When patients undergo colonoscopy, lesions are identified, removed and sent for pathological evaluation. Because of this, much of what we know about precursor lesions is based on colonoscopy studies,[2] including age- and sex-specific prevalence, multiplicity, and size distributions. Colonoscopy sensitivity to detect adenomas conditional on their size is also an important parameter in model-based CRC screening effectiveness analyses.[3]

Colonoscopy sensitivity has been informed by tandem colonoscopy studies wherein two colonoscopies are performed on the same patient. Lesions are removed at the first exam. These studies estimate the adenoma miss rate (AMR) by counting the number of adenomas missed by the first colonoscopy and found in the second exam. Meta-analyses of miss rates from tandem colonoscopy studies provide population-level adenoma miss rate estimates by adenoma size, histology and morphology.[4–6]

If one makes the assumption that the second, enhanced colonoscopy is *perfect* across all the endoscopists who participated in these studies and across all adenoma subgroups, then 1 − AMR from tandem colonoscopy studies can be interpreted as the sensitivity of the first, white-light colonoscopy (WLC). However, this strong assumption is problematic, especially for small adenomas (i.e., 1–5 mm adenomas) or flat adenomas, which have documented miss rates of 36% and 50%, respectively.[6]

To address this problem, we develop two approaches to estimate colonoscopy sensitivity from tandem colonoscopy meta-analysis while relaxing the assumption that enhanced colonoscopy is perfect. The first approach is a Bayesian hierarchical model that pools information across tandem colonoscopy studies and estimates colonoscopy sensitivity while recognizing that the second colonoscopy, while potentially improved, can also be imperfect. The second draws its motivation from the partial identification literature [7, 8] and derives an equation that expresses WLC sensitivity as a function of the adenoma miss rate and an assumed detection odds ratio of the enhanced colonoscopy versus WLC. This equation can then be used to construct an interval bounding WLC sensitivity without fitting a Bayesian model. We apply both approaches to estimate colonoscopy sensitivity using data from a recent tandem colonoscopy meta-analysis.[6] We then assess the robustness of sensitivity estimates to specification of the prior distribution that is tied to sensitivity of the second exam, as well as the agreement between the two approaches.

## 2 Methods

### 2.1 Data

Jahn et al. [6] pool sixteen randomized tandem trials, each comparing standard WLC against an enhanced, presumably more sensitive colonoscopy. They estimate and report adenoma miss rates overall and within ten subgroups (three size bands, two histology groups, two location groups, two morphology groups and adenomas overall). For each study-subgroup *j* we observe *n*_1*j*_ adenomas found at the first WLC exam and *n*_2*j*_ adenomas missed by WLC but found using enhanced colonoscopy, with *m*_*j*_ = *n*_1*j*_ + *n*_2*j*_ the total detected. The adenoma miss rate is AMR_*j*_ = *n*_2*j*_*/m*_*j*_. We estimate sensitivity from these sixteen trials, though both approaches apply to any set of tandem colonoscopy studies.

### 2.2 Model

#### 2.2.1 Data-generation process

We model each study-subgroup’s counts independently, pooling information across studies for each subgroup, with the multinomial process implied by two exams with potentially different sensitivity. The unobserved total number of adenomas present in study-subgroup *j* is *N*_*j*_ = *n*_1*j*_ + *n*_2*j*_ + *n*_miss,*j*_, where the number of adenomas missed by both exams *n*_miss,*j*_ is unknown; only the detected count *m*_*j*_ = *n*_1*j*_ + *n*_2*j*_ is observed. Study *j*’s first- and second-exam sensitivities *s*_1*j*_ and *s*_2*j*_ are defined using a non-centered hierarchical parameterization [9, 10] where

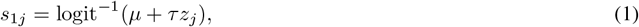

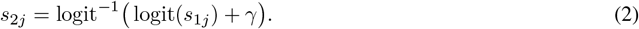

The population first-exam sensitivity is *s*_pop_ = logit^−1^(*µ*), *τ* is its between-study variation on the logit scale, the *z*_*j*_ are standardized study deviations that capture between-study variability in first-exam sensitivity, and *γ* ties the second exam to the first on the logit scale. This specification assumes the same additive increase on the logit scale across studies. Because *γ* is a difference of logits, *e*^*γ*^ is the odds ratio of detection at the second (enhanced) exam relative to the first. Setting *γ* = 0 makes both sensitivities the same.

Outcomes from the *j*th study are modeled using a multinomial distribution that describes outcomes for the *N*_*j*_ adenomas: *n*_1*j*_ found at the first exam with probability *s*_1*j*_; *n*_2*j*_ missed at the first exam and found at the enhanced exam with probability (1 *− s*_1*j*_)*s*_2*j*_; and *n*_miss,*j*_ missed by both exams with probability (1 *− s*_1*j*_)(1 *− s*_2*j*_), whose expected counts are given in SA 1.1.1. Only *n*_1*j*_ and *n*_2*j*_ are observed:

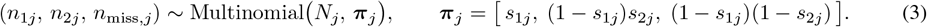

Because *N*_*j*_ is not observed, we fit the likelihood implied by Eq. 3 conditional on *m*_*j*_, which is binomial and does not involve *N*_*j*_ (SA 1.2).

#### 2.2.2 Prior Distributions

We place non-informative prior distributions on the population first-exam sensitivity *s*_pop_, its between-study variation *τ*, the standardized study deviations *z*_*j*_. Because we do not expect the data to constrain *γ*, we place an informative distribution on it and evaluate the robustness of our estimates to that prior distribution in sensitivity analyses. Our prior distributions are:

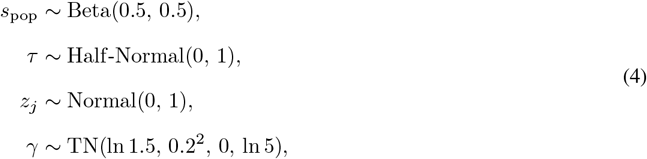

where *γ ∼* TN(*µ, σ*^2^, *a, b*) denotes a truncated Normal distribution in which *µ* and *σ*^2^ are the mean and variance of the parent Normal distribution and *a* and *b* are the lower and upper truncation bounds, respectively. We set the variance so that the lower bound of the distribution is approximately 1, reaching our lower bound that assumes WLC to be no inferior to enhanced colonoscopy (i.e., *e*^ln 1.5−1.96*×*0.2^ *≈* 1). Truncating *γ* to [0, ln 5] implies that the enhanced second exam is no worse than WLC and *at most* fivefold better on the odds scale ( 1 *≤ e*^*γ*^ *≤* 5). This is a generous upper limit for the relative performance of enhanced colonoscopy because 5 exceeds the detection odds ratios supported by enhanced-technology trials.[11]

Because we expect the prior distribution for *γ* to be informative about *s*_1_, we examined a range of different priors in two ways: We first shift the mean of the truncated Normal distribution, and we refer to the resulting specifications as Low, Baseline, High OR. The Baseline OR specification is the one in Eq. 4, with *γ ∼* TN(ln 1.5, 0.2^2^, 0, ln 5) because 1.5 is the order of magnitude of the relative OR of enhanced colonoscopy versus WLC observed in adenoma detection rate trials.[11] Sensitivity analyses explore the Low OR specification, *γ ∼* TN(ln 1, 0.2^2^, 0, ln 5), which is a lower bound if one assumes that enhanced colonoscopy is no worse than WLC, and the High OR specification, *γ ∼* TN(ln 3, 0.2^2^, 0, ln 5), as an upper bound. Finally, we present an Uninformative OR specification placing a flat prior distribution on *γ, γ ∼* Uniform[0, ln 5]. This last specification encompasses the full range of sensitivity estimates that one can obtain from this model given this range of ORs from 1 to 5. Therefore, these alternative prior specifications are meant to cover the full plausible range of *γ* and therefore of *s*_1_ and *s*_2_. Together, these prior distributions imply prior distributions on *s*_1_, *s*_2_, AMR and *e*^*γ*^; Table S1 reports their prior means and 95% intervals next to the posterior estimates.

#### 2.2.3 Statistical Analyses

We fit the model in Stan[12] using Hamiltonian Monte Carlo estimation.[13] Models were estimated for each lesion subgroup using four chains of 4,000 iterations (2,000 warmup). Convergence was assessed with the potential scale reduction factor 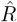, effective sample size, and the number of divergent transitions.[14] Across every fit reported here, 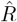 did not exceed 1.0053, the smallest effective sample size was 1,482, and there were 0 divergent transitions.

#### 2.2.4 Posterior Estimates

For each of the ten lesion subgroups, we report the posterior mean and 95% credible interval of the first-exam (white-light) sensitivity *s*_1*j*_, the second-exam (enhanced) sensitivity *s*_2*j*_, the model-implied AMR, and the detection odds ratio *e*^*γ*^ comparing WLC and enhanced colonoscopy.

We also quantify the extent to which assuming that the second exam is perfect can bias sensitivity estimates, defining this bias as Bias = (1 − AMR) *− s*_1_. To check that our estimates are consistent with the published evidence, we compare our model-implied miss rate to previously reported pooled miss rates estimated using a random-effects meta-analysis.[6]

#### 2.2.5 Sensitivity Analyses

To reveal how the identifying prior distribution affects sensitivity estimates, we fit the model under the four specifications for the prior distribution of *γ* described above (Eq. 4): Low OR, with parent mean ln 1, the point at which the two exams are equally sensitive; Baseline OR, with parent mean ln 1.5; High OR, with parent mean ln 3, larger than any pooled advantage the trial literature reports; and Uninformative OR, the flat prior distribution over [0, ln 5]. To confirm that the prior distributions imply plausible data before conditioning on the observations, we performed prior predictive checks by drawing from the prior distributions with the likelihood switched off, reported alongside the posterior in Table S1.

### 2.3 A frequentist approach to bounding *s*_1_

WLC sensitivity *s*_1_, AMR, and the detection odds ratio of the enhanced exam relative to WLC, *OR* = *e*^*γ*^ are related by this equation (derivation in SA 1.1):

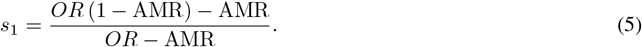

This formula expresses WLC sensitivity in terms of the true AMR and *OR*, so it allows estimation of WLC sensitivity from an estimate 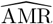 and an assumed *OR*. When the second exam is perfect, *OR → ∞* and *s*_1_ = 1 − AMR. If one assumes that there is no difference between the tests, then *OR* = 1. If one assumes that the enhanced exam is better than WLC, *OR* can be set to a suitable value, ideally informed by other studies or expert opinion. For a fixed *OR, s*_1_(*OR, ·*) is a continuous, strictly decreasing function of AMR. We can carry the sampling uncertainty in 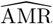 through Eq. 5 analytically (SA 1.1.9) to compute a confidence interval for *s*_1_ from the 95% confidence bounds of 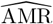, [AMR_*L*_, AMR_*U*_ ]:

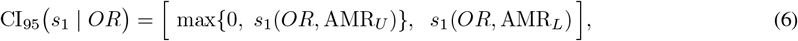

The truncation at zero handles the case in which *s*_1_(*OR*, AMR) falls below zero, which occurs when AMR *> OR/*(1 + *OR*) and the iso-*OR* curve no longer crosses the iso-AMR curve (SA 1.1.10, Figure S1).

When the *OR* is not known but is assumed to lie in a range [*OR*_*L*_, *OR*_*U*_ ], *s*_1_ is partially identified:[7] the data and the assumption are consistent with a set of values of *s*_1_ rather than a single value. In this case we can estimate a plausible range for *s*_1_, but not a confidence interval; the plausible range does not carry the probabilistic or frequentist interpretation of a credible or confidence interval. For each *OR* value, we can estimate the 95% CI using Eq. 6. We set the lower bound of the plausible range to min_*OR*_ max*{*0, *s*_1_(*OR*, AMR_*U*_ )*}* and the upper bound to max_*OR*_ *s*_1_(*OR*, AMR_*L*_), which is equivalent to pairing the smallest *OR* with the largest miss rate and the largest *OR* with the smallest miss rate. If we assume that the enhanced exam is no worse than WLC at detecting adenomas, the lower bound for *OR* is 1. Further, we can choose a high value for *OR* that exceeds estimates found by available evidence for the advantage of enhanced technologies. Taking *OR*_*L*_ = 1 and *OR*_*U*_ = 5 (an advantage larger than reported in the literature [11]), the two bounds of the plausible range become

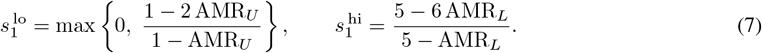

This plausible range reflects two sources of uncertainty: fixing *OR* and widening AMR to its confidence limits isolates sampling error, while fixing AMR at its point estimate and sweeping over *OR* values reflects uncertainty about the relative performance of the tests. We evaluate Eq. 6 and Eq. 7 for each of the ten subgroups, taking AMR and its confidence interval from the Jahn et al. 2025 [6] random-effects meta-analysis. This approach can be applied to any published tandem colonoscopy meta-analysis and does not require anything other than a range for *OR* and a confidence interval for AMR.

### 2.4 External evidence on detection odds ratios

Both the Bayesian and the frequentist approach we propose depend on assumptions about the detection odds ratio *OR* of the enhanced colonoscopy compared with WLC. This assumption can be informed by trials that compare enhanced technologies against WLC. A meta-analysis of these trials reported pooled odds ratios for adenoma and polyp detection of 1.35 to 1.51 in favor of the enhanced exam.[11] Our baseline specification uses a prior distribution for *γ* allowing it to be uncertain while placing the mean of its parent Normal distribution at ln 1.5 (Eq. 4). We recognize that these trials measure detection rates per patient rather than sensitivity per lesion. Although person-level detection rates are not the same as the sensitivity, here we let the order of magnitude of the detection odds ratio of enhanced colonoscopy vs WLC inform our priors for the odds ratio of sensitivity. Throughout this paper we confine this odds ratio to be at least 1 (i.e., enhanced colonoscopy is no worse than WLC) and 5, which is well above the upper bound reported by [11].

## 3 Results

### 3.1 Hierarchical Bayesian model estimates

#### 3.1.1 Sensitivity estimates in the baseline specification

Under the Baseline OR specification, the estimated overall first-exam sensitivity of WLC is 0.54 [0.41, 0.63] (posterior mean and 95% credible interval), below the conventional 1 − AMR value of 0.65 [0.60, 0.70] (Table 1). First-exam sensitivity increases with lesion size, from 0.49 [0.35, 0.60] for 1–5 mm adenomas to 0.64 [0.43, 0.78] for 6–9 mm and 0.84 [0.70, 0.93] for adenomas *≥* 10 mm, and is lowest for flat lesions (0.28 [0.05, 0.58]). The second-exam (enhanced) overall sensitivity was 0.62 [0.44, 0.77]. Our model-implied miss rate (overall 0.35 [0.30, 0.40]) aligns well with the previously reported miss rate estimated by Jahn et al. [6] (overall 0.34 [0.30, 0.38]; Table 2), both overall and across subgroups (Figure 1). The Bayesian and meta-analytic estimates of 1 − AMR nearly coincide, while the first-exam sensitivity *s*_1_ is less than both estimates. As expected, the joint posterior of *s*_1_ and *s*_2_ lies along a ridge where AMR is constant across prior distribution specifications, and above the *s*_1_ = *s*_2_ line by construction (Figure S2).

**Table 1:** Posterior estimates by subgroup under the Baseline OR specification.

|  | Sensitivity |  | AMR | Bias | Enhanced vs WLC OR |
| --- | --- | --- | --- | --- | --- |
|  | First exam | Second exam |  |  |  |
| Overall | 0.54 [0.41, 0.63] | 0.62 [0.44, 0.77] | 0.35 [0.30, 0.40] | 0.12 [0.06, 0.20] | 1.46 [1.04, 2.12] |
| 1-5 mm | 0.49 [0.35, 0.60] | 0.58 [0.38, 0.74] | 0.37 [0.32, 0.44] | 0.14 [0.07, 0.23] | 1.49 [1.04, 2.16] |
| 6-9 mm | 0.64 [0.43, 0.78] | 0.72 [0.49, 0.86] | 0.29 [0.19, 0.41] | 0.07 [0.03, 0.18] | 1.51 [1.06, 2.18] |
| >= 10 mm | 0.84 [0.70, 0.93] | 0.89 [0.76, 0.96] | 0.14 [0.07, 0.25] | 0.02 [0.00, 0.05] | 1.53 [1.06, 2.24] |
| Non-advanced | 0.39 [0.23, 0.51] | 0.48 [0.24, 0.68] | 0.42 [0.38, 0.47] | 0.19 [0.10, 0.32] | 1.46 [1.04, 2.11] |
| Advanced | 0.75 [0.58, 0.87] | 0.81 [0.64, 0.92] | 0.21 [0.12, 0.32] | 0.04 [0.01, 0.10] | 1.51 [1.06, 2.16] |
| Proximal | 0.50 [0.33, 0.63] | 0.59 [0.37, 0.76] | 0.37 [0.30, 0.45] | 0.13 [0.06, 0.24] | 1.51 [1.06, 2.17] |
| Distal | 0.51 [0.37, 0.62] | 0.60 [0.40, 0.76] | 0.36 [0.30, 0.42] | 0.13 [0.06, 0.22] | 1.48 [1.05, 2.14] |
| Polypoid | 0.64 [0.44, 0.79] | 0.72 [0.51, 0.87] | 0.28 [0.19, 0.40] | 0.07 [0.02, 0.17] | 1.52 [1.08, 2.19] |
| Flat | 0.28 [0.05, 0.58] | 0.40 [0.08, 0.73] | 0.51 [0.34, 0.63] | 0.21 [0.08, 0.36] | 1.84 [1.35, 2.51] |

*Notes:* First- and second-exam sensitivity are the posterior white-light ( $s_1$ ) and enhanced ( $s_2$ ) detection probabilities; AMR is the model-implied adenoma miss rate computed from the posterior; bias is $\text{Bias} = (1 - \text{AMR}) - s_1$ , the amount by which a naive sensitivity estimate $1 - \text{AMR}$ (which treats the enhanced pass as perfect) overstates first-exam sensitivity; enhanced vs. WLC OR is the detection odds ratio $e^\gamma$ of the enhanced colonoscopy vs WLC. Values are posterior mean [95% credible interval].

**Table 2:** WLC sensitivity bounds implied by estimated adenoma miss rates in Jahn 2025.

|  | AMR | First-exam sensitivity at a fixed OR |  |  | OR from<br>1 to 5 |
| --- | --- | --- | --- | --- | --- |
|  |  | OR = 1 | OR = 1.5 | OR = 5 |  |
| Overall | 0.34 [0.30, 0.38] | 0.49 [0.38, 0.57] | 0.56 [0.49, 0.63] | 0.64 [0.59, 0.68] | 0.38 to 0.68 |
| 1-5 mm | 0.36 [0.33, 0.40] | 0.43 [0.34, 0.50] | 0.52 [0.46, 0.58] | 0.61 [0.57, 0.65] | 0.34 to 0.65 |
| 6-9 mm | 0.27 [0.20, 0.36] | 0.62 [0.45, 0.74] | 0.67 [0.53, 0.76] | 0.71 [0.62, 0.79] | 0.45 to 0.79 |
| $\geq 10$ mm | 0.12 [0.07, 0.20] | 0.86 [0.76, 0.92] | 0.87 [0.77, 0.92] | 0.87 [0.80, 0.92] | 0.76 to 0.92 |
| Non-advanced | 0.42 [0.38, 0.45] | 0.29 [0.17, 0.39] | 0.42 [0.35, 0.49] | 0.55 [0.50, 0.59] | 0.17 to 0.59 |
| Advanced | 0.21 [0.14, 0.29] | 0.74 [0.60, 0.83] | 0.76 [0.64, 0.84] | 0.78 [0.69, 0.85] | 0.60 to 0.85 |
| Proximal | 0.36 [0.30, 0.42] | 0.45 [0.28, 0.57] | 0.53 [0.42, 0.63] | 0.62 [0.54, 0.68] | 0.28 to 0.68 |
| Distal | 0.36 [0.31, 0.40] | 0.45 [0.32, 0.55] | 0.53 [0.45, 0.61] | 0.62 [0.56, 0.67] | 0.32 to 0.67 |
| Polypoid | 0.27 [0.20, 0.36] | 0.63 [0.45, 0.75] | 0.67 [0.53, 0.77] | 0.71 [0.62, 0.79] | 0.45 to 0.79 |
| Flat | 0.50 [0.36, 0.64] | 0.01 [0.00, 0.45] | 0.25 [0.00, 0.53] | 0.45 [0.26, 0.62] | 0.00 to 0.62 |

Notes: AMR is the random-effects pooled adenoma miss rate for the subgroup, estimate [95% confidence interval], reproduced from the tandem trials of Jahn et al. [6] that report that subgroup: all sixteen for the overall row, and between six and eleven for the remaining rows. The three fixed-OR columns give $s_1(OR, AMR)$ (Eq. 5) at the pooled estimate, with the 95% confidence interval obtained by transforming the AMR confidence limits and reversing them (Eq. 6). The last column is the union of those intervals over $OR \in [1, 5]$ (Eq. 7), running from the $OR = 1$ lower limit to the $OR = 5$ upper limit. Values of 0.00 appear whenever $AMR > OR/(1 + OR)$ because at that point the odds-ratio assumption no longer provides a lower bound for $s_1$ .

**Figure 1:**
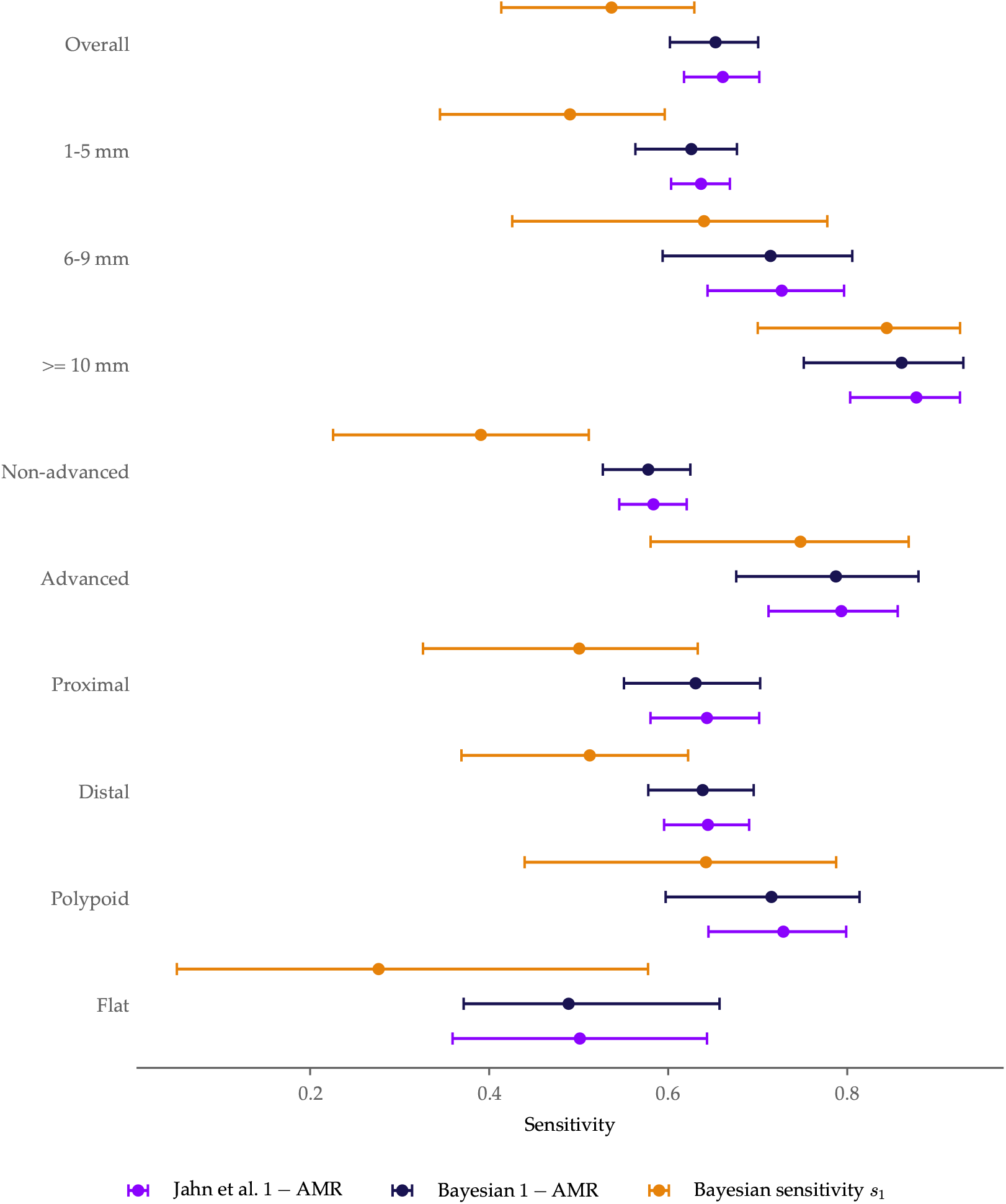
Sensitivity and AMR estimates. *Notes:* Per-subgroup forest plot on the sensitivity scale, Baseline OR specification: Jahn et al. 1 − AMR, Bayesian 1− AMR, and Bayesian first-exam (white-light) sensitivity *s*_1_. The Jahn et al. series is our reproduction of their random-effects pooled miss rate on the same trial counts, rather than their published figures. Points are estimates and bounds are the 95% confidence interval for the random-effects estimate and 95% posterior credible intervals for the Bayesian estimates.

#### 3.1.2 Sensitivity Analyses

Sensitivity analyses that examined different prior distributions for *γ* show how much the results depend on the prior distribution. The estimated first-exam sensitivity depends on the assumed detection odds ratio (Figure S3, Table S1). Overall *s*_1_ rises from 0.49 [0.37, 0.59] under Low OR to 0.54 [0.41, 0.63] under Baseline OR and 0.61 [0.54, 0.67] under High OR (posterior mean and 95% credible interval). Under Uninformative OR we estimate *s*_1_ at 0.56 [0.41, 0.66]. A larger detection advantage implies fewer adenomas missed by both exams and so a higher first-exam sensitivity for the same observed counts. The posterior densities of *s*_1_ vary across the four specifications (Figure S4), reflecting that the data alone identify the miss rate but not each exam’s sensitivity separately, and that sensitivity estimates depend heavily on the prior distribution of *γ*. The differences between the posterior distributions across the prior specifications are wider for non-advanced and flat lesions, whose high AMRs mean that a long stretch of the iso-AMR ridge lies above the *s*_1_ = *s*_2_ line. Conversely, the differences between the posterior distributions are narrowest for adenomas *≥* 10 mm, which have low AMRs. In contrast, the model-implied miss rate is stable across specifications: the posteriors of 1 − AMR under all four nearly coincide (Figure S5), because the data identify the AMR regardless of the assumed *OR*.

#### 3.1.3 Bias implied by alternative specifications

The gap between the conventional and Bayesian estimates of first-exam sensitivity, Bias = (1 − AMR) *− s*_1_, is the amount by which treating the enhanced exam as perfect overstates WLC sensitivity. It shrinks as the assumed detection odds ratio grows, since 1 − AMR is stable while *s*_1_ rises, and is zero only in the limit of a perfect enhanced exam. Under the Baseline OR specification, the estimated bias in 1 − AMR as an estimate of sensitivity is 0.12 [0.06, 0.20] (posterior mean and 95% credible interval) (Table 1). The practical consequence is that bias is greatest in the subgroups with the highest miss rates, non-advanced and flat lesions, and smallest for adenomas *≥* 10 mm (Table 1).

### 3.2 Frequentist bounds on WLC sensitivity

Table 2 evaluates Eq. 6 and Eq. 7 for each of the ten subgroups, using the adenoma miss rates estimated by Jahn et al. [6]. Overall, the plausible range for WLC sensitivity is 0.38 to 0.68. The plausible range narrows with lesion size to 0.76 to 0.92 for adenomas *≥* 10 mm, and is widest where the miss rate is high (e.g., 0.17 to 0.59 for non-advanced adenomas). For adenomas *≥* 10 mm the three fixed-*OR* intervals are nearly identical (the sampling error in AMR accounts for almost the entire range), whereas for non-advanced adenomas each fixed-*OR* interval is narrow and the range is driven by the choice of *OR*.

The overall Bayesian estimate under the baseline specification, 0.54 [0.41, 0.63], falls within the 0.38 to 0.68 frequentist plausible range based on varying *OR* from 1 to 5. Both approaches place overall WLC sensitivity below the conventional 1 − AMR estimate, and both suggest lower sensitivity for small, non-advanced, and flat lesions.

## 4 Discussion

Tandem colonoscopy data identify the adenoma miss rate well, but they do not identify the sensitivity of either exam without an external assumption about the sensitivity of enhanced colonoscopy. The approaches developed in this paper make that assumption explicit. Under our Baseline OR specification, overall WLC sensitivity was 0.54 [0.41, 0.63], about twelve percentage points below the conventional 1 − AMR estimate (bias 0.12 [0.06, 0.20]); the frequentist plausible range, allowing *OR* from 1 to 5, was 0.38 to 0.68. Both frequentist and Bayesian approaches estimated lower sensitivity when miss rates are highest. These colonoscopy sensitivity estimates are lower than the colonoscopy sensitivity values that have been used in CRC screening decision analyses.[1, 15, 16]

We allowed a range of enhanced colonoscopy differences that were greater than estimated improvements. In back-to-back studies, the second exam can have higher sensitivity relative to the first because of enhancements such as the use of narrow band imaging or AI technology,[17] and also because patients undergoing tandem colonoscopy are likely to have better preparation and deeper sedation than colonoscopy in practice[18] and the field is primed for better detection of lesions based on anatomic position and straightening of folds in the large intestine. Our model allows for this inherent feature of back-to-back studies by allowing the sensitivity of the enhanced colonoscopy study to be better than the sensitivity of the first colonoscopy. Our model also used random effects to allow for between-study heterogeneity due to differences in bowel preparation, endoscopist experience, withdrawal technique, procedural indication, and patient population.[5]

Our analyses demonstrate that overall sensitivity estimates are driven by the sensitivity to detect small, 1–5 mm, lesions. This is due to the relatively large number of small lesions,[19] and explains how colonoscopy can be a highly effective way to prevent colorectal cancer even if the sensitivity to detect small lesions is much lower than previously believed.[20] These small lesions rarely transition to colorectal cancer,[19] and repeated screening offers the opportunity for detection as they grow.

Colonoscopy sensitivity assumptions have implications beyond colonoscopy screening itself. Because every non-invasive screening strategy relies on follow-up colonoscopy, WLC sensitivity limits the projected effectiveness of stool- and blood-based screening as well;[21] Jahn et al. [6] make the related point that treating WLC as a gold standard inflates the apparent sensitivity of the tests it is used to validate. Colonoscopy sensitivity assumptions also impact projected absolute benefits of colonoscopy screening, though the ranking of strategies may be robust to these assumptions.[3] Modelers should adopt more realistic sensitivity estimates. We recommend the estimates from our baseline Bayesian specification. When new tandem colonoscopy data become available, the frequentist approach we present also provides a way to obtain plausible ranges for sensitivity given an assumption about the odds ratio of the enhanced test.

## Data Availability

Data analyzed in this study were extracted from a previously published tandem colonoscopy meta-analysis. The analysis code is available from the corresponding author on reasonable request.

## Author contributions

P.N.L. conceived the study, developed the statistical model, performed the analyses, and wrote the manuscript. R.B.I. reviewed and edited the manuscript. C.M.R. reviewed the methods and contributed to the manuscript. All authors read and approved the final manuscript.

## Supplementary Materials

### SA 1 Supplementary Methods

#### SA 1.1 Asymptotic relationships

This section works out algebraic relationships between the expectations of key model parameters. Those relationships build intuition about the behavior of this model and lead to an identification argument: in the setting of tandem colonoscopy trials, an assumption about the relative performance of the second, enhanced colonoscopy (EC) exam identifies the sensitivity of white-light colonoscopy (WLC). Throughout this section, we drop the study subscript *j* and write AMR for the expected adenoma miss rate and *α* for the expected number of adenomas missed by both exams per adenoma detected.

#### SA 1.1.1 Expected counts

Of *N* adenomas present in the study-subgroup population, WLC finds a fraction with sensitivity *s*_1_. Of the adenomas not found by WLC, EC finds a fraction with sensitivity *s*_2_:

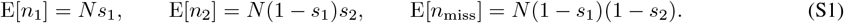

Therefore, the expected number of detected adenomas is E[*m*] = *N* [*s*_1_ + (1 *− s*_1_)*s*_2_].

##### SA 1.1.2 AMR and sensitivity

In expectation, the adenoma miss rate AMR = *n*_2_*/*(*n*_1_ + *n*_2_) is related to sensitivity. Substituting Eq. S1 and canceling *N*,

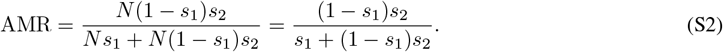

This relationship between AMR and *s*_1_ and *s*_2_ is useful because AMR is well identified by these two observed quantities *n*_1_ and *n*_2_ without depending on *N* and yet it constrains the values that *s*_1_ and *s*_2_ can take.

##### SA 1.1.3 WLC sensitivity when EC is perfect

If the enhanced colonoscopy sensitivity is perfect (*s*_2_ = 1), then AMR = 1 *− s*_1_, hence *s*_1_ = 1 − AMR. We show below that this assumption can be relaxed and that doing so yields a lower bound for *s*_1_.

##### SA 1.1.4 Double miss rate *α* and sensitivity

Let *α* be the fraction of adenomas never removed (i.e., missed by both exams) relative to the number of adenomas detected, a “double” miss rate. If *n*_miss_ is the number of adenomas missed by both exams, *α* = *n*_miss_*/*(*n*_1_ + *n*_2_). As above, *n*_miss_ = *N* (1 *− s*_1_)(1 *− s*_2_) and *n*_1_ + *n*_2_ = *N* [*s*_1_ + (1 *− s*_1_)*s*_2_]. Therefore *α* can also be defined independent of *N* as

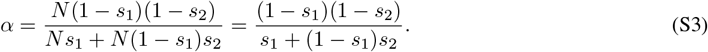

Whereas AMR is the fraction of adenomas missed by the first exam relative to those detected, *α* is the fraction missed by both exams relative to those detected by either exam.

##### SA 1.1.5 Expected counts per adenoma detected

For the purposes of this derivation, it can be useful to express the expected adenoma counts by expected number of adenomas detected *m* = *n*_1_ + *n*_2_ rather than by *N* . For instance, given the relationships above, the expected number of adenomas by group per adenoma detected are:

| Group | Expected counts per adenoma detected |
| --- | --- |
| Found at first exam | $1 - \text{AMR}$ |
| Missed at first, found at second | $\text{AMR}$ |
| Missed at both exams | $\alpha$ |
| Detected | $1$ |
| Missed by the first exam | $\text{AMR} + \alpha$ |
| Present | $1 + \alpha$ |

Given these expectations, we can express each sensitivity as a ratio of two rows in this table:

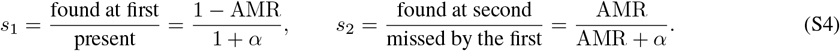

It should be clearer now why *α* is important for *s*_1_. If *α* = 0, then *s*_1_ = 1 − AMR.

##### SA 1.1.6 Detection odds and odds ratios of EC vs WLC

We now have an expression for *s*_1_ as a function of the expected *α*. That does not take us far on its own, because it is hard to make a claim about *α* a priori. What we do know is that the enhanced colonoscopy should be *no worse* than white-light colonoscopy at detecting adenomas. That claim imposes a second relationship between *s*_1_ and *s*_2_, and so aids identification. We call it an *identification assumption*: enhanced colonoscopy is no worse than white-light colonoscopy.

We will do this algebraically by defining the odds of detection for the first and second exam *o*_1_ and *o*_2_

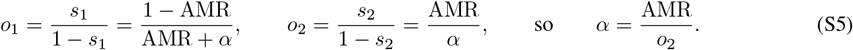

This relationship makes sense because *α →* 0 only when *o*_2_ *→ ∞*. That limit is extreme, yet it is exactly what treating the second exam as perfect assumes.

##### SA 1.1.7 Double miss rate *α* and the detection odds ratio OR

Let *OR* = *o*_2_*/o*_1_ be the detection odds ratio of the enhanced exam relative to white-light colonoscopy. We want an expression that relates *α* to AMR and *OR*. Substituting Eq. S5,

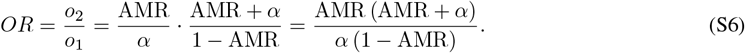

Solving for *α*:

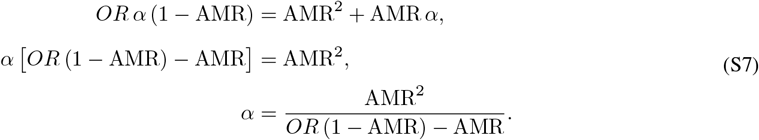

The implied detection odds of the enhanced exam follow from Eq. S5,

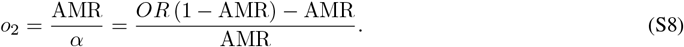

##### SA 1.1.8 WLC sensitivity *s*_1_ as a function of OR and AMR

Now that we defined relationships between *α*, AMR, and *OR*, we are ready to use those to work towards trying to bound *s*_1_ given external information about *OR*. So far, because the data alone only identifies AMR well, we can see in eqs. S2 and S4 that placing no constraint in *α* allows multiple pairs (*s*_1_, *s*_2_) to result in the same AMR. Those pairs form a curve in the (*s*_1_, *s*_2_) plane (see *iso-AMR* curves in Figure S1). Those pairs are obtained by solving Eq. S2 for *s*_2_:

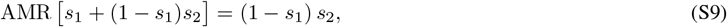

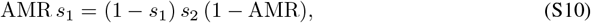

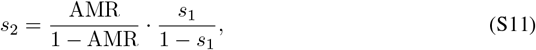

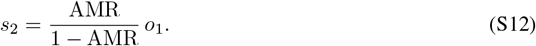

A second family of curves follows from the definition of the detection odds ratio, *OR* = *o*_2_*/o*_1_ (see *iso-OR* curves in Figure S1). Rearranging and converting the odds back to a probability gives the *iso-OR* curve, the sensitivity pairs whose detection odds ratio equals a given *OR*:

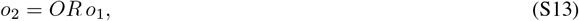

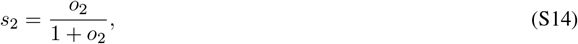

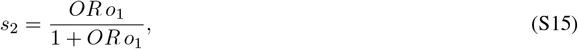

substituting *o*_1_ = *s*_1_*/*(1 *− s*_1_), *s*_2_ can be expressed as:

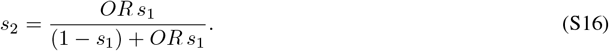

**Figure S1:**
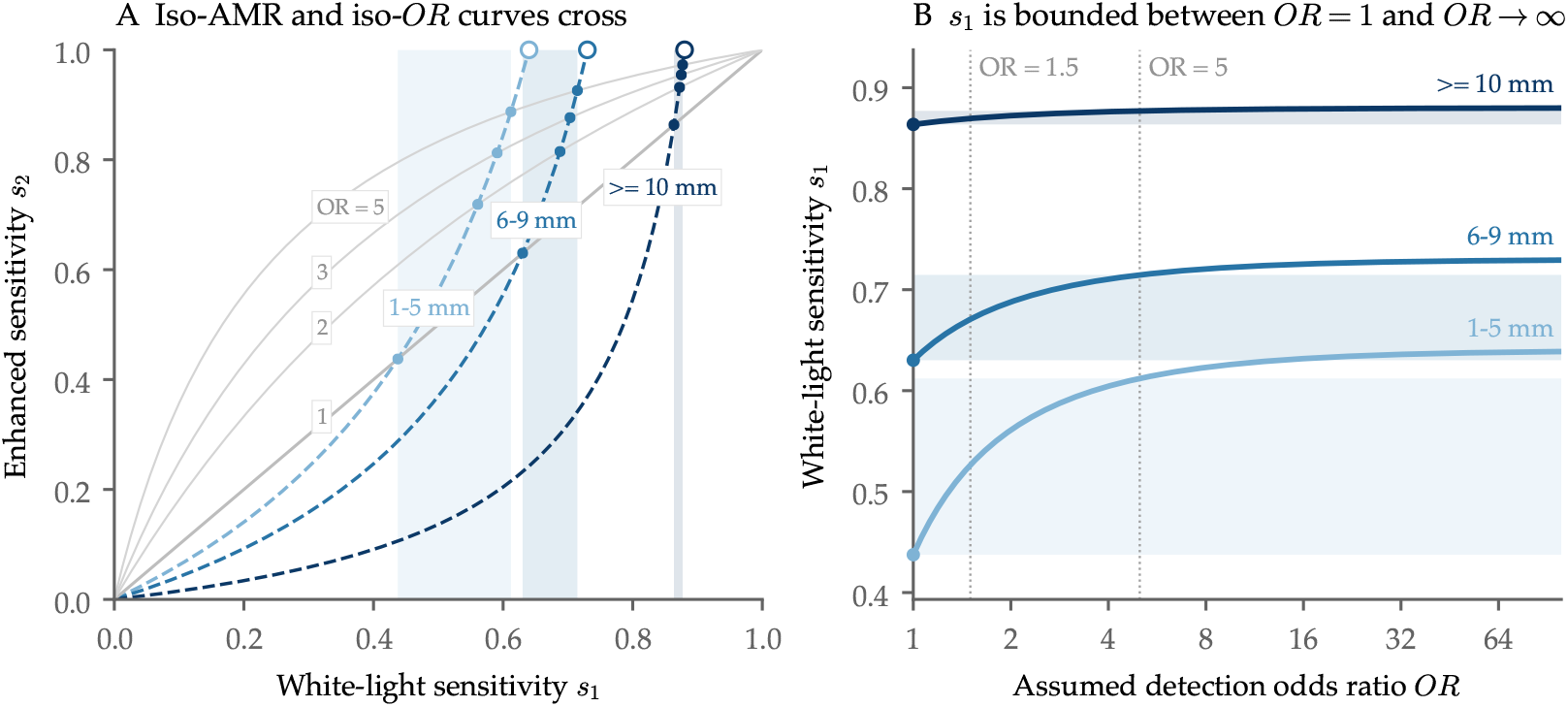
Relationships between *s*_1_ and *s*_2_. *Notes:* (**A)** Iso-AMR curves (dashed, one per size band, Eq. S12) cross iso-OR curves (faint grey, OR = 1, 2, 3, 5, Eq. S16) at the sensitivity pair an assumed odds ratio selects (dots, Eq. S25); open circles are the OR→ ∞ limit *s*_1_ = 1 − AMR. (**B)** First-exam sensitivity at that crossing against the assumed odds ratio, by size band; dotted lines mark OR = 1.5 and OR = 5.

Both the iso-AMR curves and the iso-OR curves pass through the origin, but their other intersection is where the odds-ratio relationship between the two sensitivities and the evidence about the adenoma miss rates is informative. We do not know *OR* precisely but if we make the assumption that *OR ≥ x*, then that helps restrict the range for the first sensitivity.

Setting Eq. S12 equal to Eq. S15 and canceling the common factor *o*_1_:

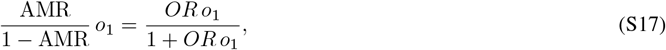

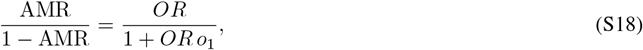

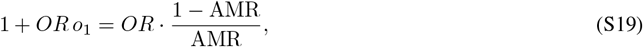

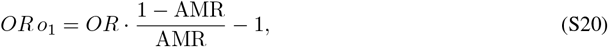

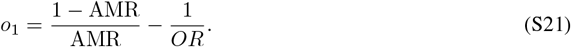

The first term is the detection odds the first exam would have if the second exam were perfect. The second term is the penalty for allowing it to be imperfect, which vanishes only as *OR → ∞*. Over a common denominator,

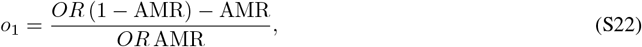

and *o*_2_ = *OR o*_1_ recovers Eq. S8. To convert Eq. S22 to a probability, note that *OR* AMR cancels against −AMR in the numerator of 1 + *o*_1_:

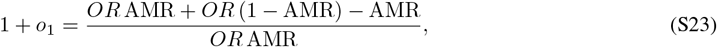

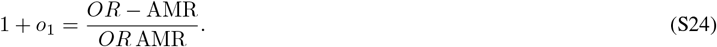

The factor *OR* AMR therefore cancels in the ratio,

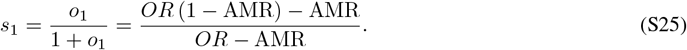

Applying the same conversion to *o*_2_ = *OR* (1 − AMR) − AMR */*AMR gives 1 + *o*_2_ = *OR* (1 − AMR)*/*AMR, so the factor AMR cancels,

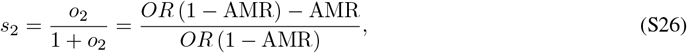

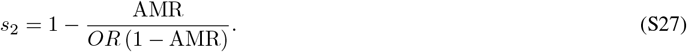

The data supply AMR, an external assumption supplies *OR*, and together they select a single point on the iso-AMR curve (Figure S1).

##### SA 1.1.9 Deriving an interval for *s*_1_ given uncertainty in AMR

All equations above hold in expectation, but tandem colonoscopy studies have finite sample sizes, so AMR is not perfectly known. Here we carry the sampling uncertainty in AMR through Eq. S25 to obtain an interval for *s*_1_, first at a fixed *OR* and then over a range of *OR*. Both steps are analytic, because *s*_1_ is monotone in each of its two arguments.

Write the crossing of Eq. S25 as a function of both the assumed odds ratio and the miss rate,

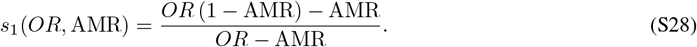

This equation recovers the fact that *s*_1_ *→* 1 − AMR when *s*_2_ *→* 1, which is the same as *OR → ∞*. To see that, one can divide the numerator and denominator by *OR*, which gives:

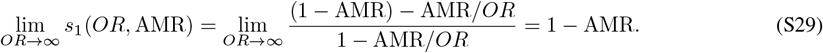

##### SA 1.1.10 An interval for *s*_1_ given a fixed *OR*

Let [AMR_*L*_, AMR_*U*_ ] be the 95% confidence interval for estimated AMR from the random-effects meta-analysis. For a fixed *OR, s*_1_(*OR, ·*) is a continuous, strictly decreasing function of AMR, so a confidence interval for *s*_1_ follows by transforming the bounds of the confidence interval and reversing their order,

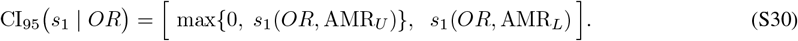

The truncation at zero in Eq. S30 handles the case in which *s*_1_(*OR*, AMR) falls below zero, which occurs when AMR *> OR/*(1 + *OR*) and the iso-*OR* curve no longer crosses the iso-AMR curve (Figure S1).

##### SA 1.1.11 An interval for *s*_1_ over a range of *OR*

If we assume that the odds ratio is within a range [*OR*_*L*_, *OR*_*U*_ ], we can also report a plausible range for *s*_1_ given the equations above. We do so by pairing the smallest *OR* with the largest miss rate, and the largest *OR* with the smallest miss rate. The union of the *OR* intervals is therefore itself an interval:

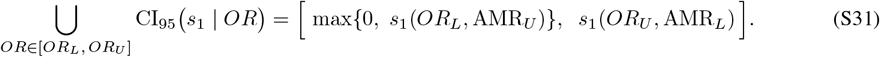

Taking *OR*_*L*_ = 1 (EC is no worse than WLC) and *OR*_*U*_ = 5, an advantage far larger than any reported in the new-technology-device trials,[11] the two endpoints become

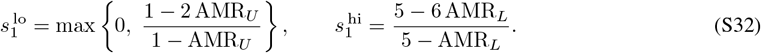

Eq. S32 is a finite-sample counterpart of the pair (1 − 2AMR)*/*(1 − AMR), 1 − AMR in Eq. S25 at *OR* = 1 and in the limit *OR → ∞*. The lower endpoint is (1 − 2AMR)*/*(1 − AMR) evaluated at the upper confidence limit for the miss rate, and the upper endpoint replaces the *OR → ∞* limit 1 − AMR_*L*_ with the upper bound of the odds ratio (here set at 5). This plausible range represents two sources of uncertainty. Fixing *OR* and widening AMR to its confidence limits isolates sampling error, while fixing AMR at its point estimate and sweeping *OR* isolates the identifying assumption. Eq. S30 and Eq. S32 correspond to Eq. 6 and Eq. 7 of the main text, which Table 2 evaluates for each of the ten subgroups.

#### SA 1.2 Likelihood function

The data-generation process in Eq. 3 is multinomial, but the likelihood we fit for each study-subgroup is binomial, modeling the number of adenomas found at the second exam given the number of adenomas detected. This section derives the binomial likelihood from the multinomial data generation process by conditioning on the number of adenomas detected *m*. We drop the study subscript *j* for clarity. As in Eq. 3, *p*_miss_ = (1 *− s*_1_)(1 *− s*_2_) is the probability that an adenoma is missed by both exams. The multinomial mass function of Eq. 3 is

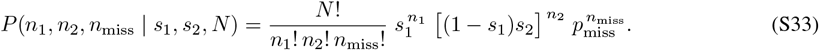

The number of adenomas missed by both exams, *n*_miss_, is not observed, so the total *N* = *m* + *n*_miss_ is unknown as well. Eq. S33 is therefore not yet a likelihood we can evaluate.

It is easier to understand our likelihood function if we generate the counts in two steps instead of one multinomial step. In the first step, each of the *N* adenomas is either detected by one of the two exams, with probability 1 *− p*_miss_ = *s*_1_ + (1 *− s*_1_)*s*_2_, or missed by both, so *m* is binomial with *N* trials and success probability 1 *− p*_miss_. In the second step, each detected adenoma is detected either by the first exam or the second exam. Among *m* detected adenomas, the probability of having been found at the second exam (only) is the adenoma miss rate:

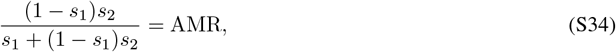

the expected miss rate of Eq. S2. So *n*_2_ is binomial with *m* trials and success probability AMR, and *n*_1_ = *m − n*_2_. Eq. S33 is therefore the product Binomial(*m* | *N*, 1 *− p*_miss_) *×* Binomial(*n*_2_ | *m*, AMR), and only the first factor involves *N* . We fit the second factor, the conditional likelihood of *n*_2_ given *m*, which does not involve *N* .

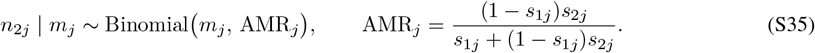

The discarded factor depends on the sensitivities only jointly with *N*, so without knowledge of *N* the number detected carries essentially no information about *s*_1_ and *s*_2_. See Sanathanan (1972)[22] for a deeper treatment of multinomial models with unknown *N* .

**Figure S2:**
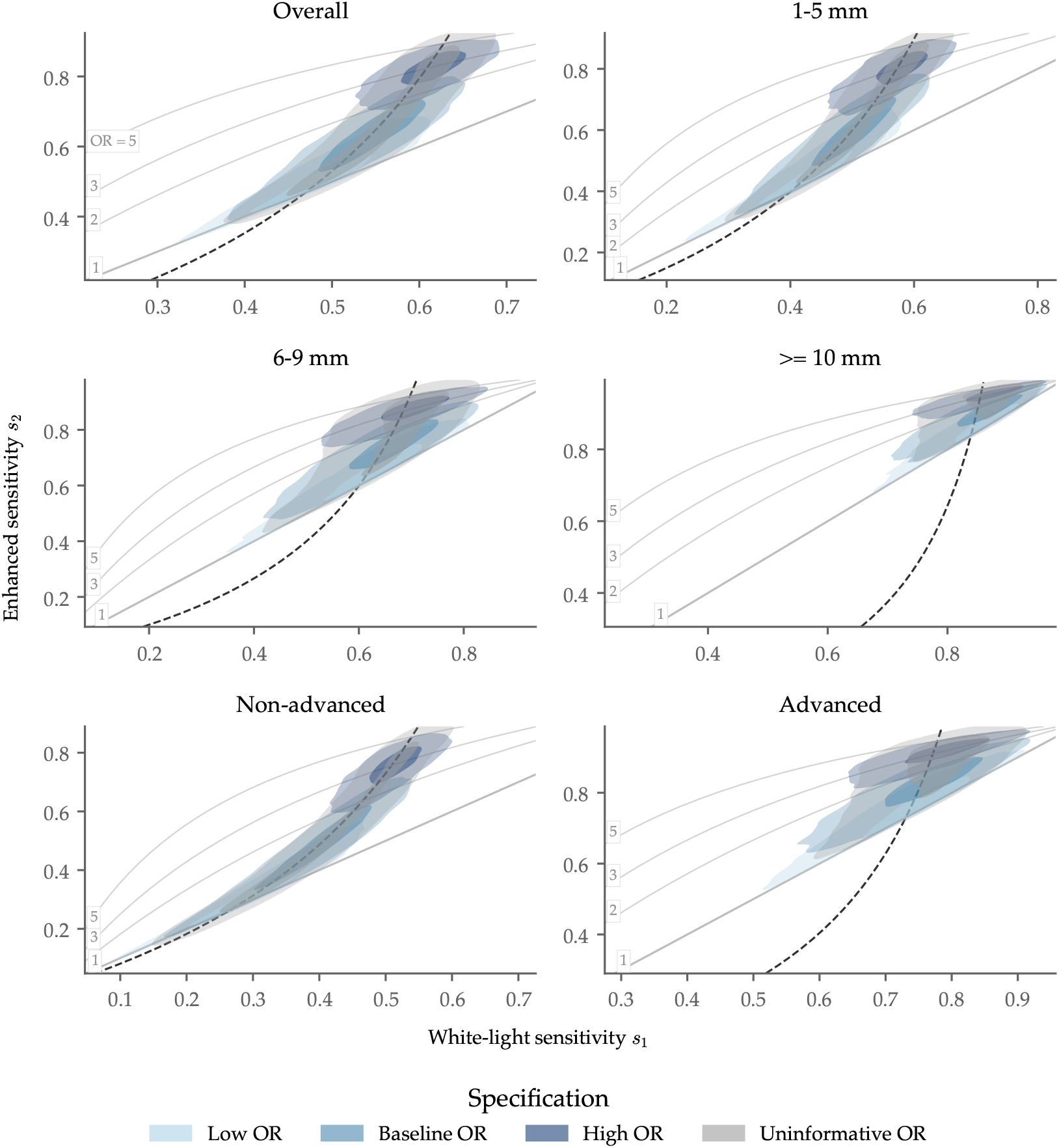
Joint posterior of white light and enhanced colonoscopy sensitivity. *Notes:* Joint posterior of first-(*s*_1_) versus second-exam (*s*_2_) sensitivity for six of the ten subgroups. Table 1 and Table S1 report all ten). Figure shows 50% and 95% highest-density regions under each of the four prior specifications, overlaid on a curve (dashed) implied by the subgroup’s estimated adenoma miss rate. The faint grey curves are the iso-OR contours (Eq. S16). The lowest of them, *OR* = 1, is the line where both exams have the same sensitivity. The four specifications differ in the prior distribution they place on *γ* (Eq. 4). The Low, Baseline and High OR place a Normal prior distribution on *γ* with mean ln 1, ln 1.5 and ln 3 and standard deviation 0.2, truncated to [0, ln 5]. A standard deviation of 0.2 was chosen because in our baseline scenario that results in the lower bound for OR to be approximately 1 (i.e., in the worst-case if our baseline distribution, enhanced colonoscopy is no worse than WLC). As an additional sensitivity analysis, we also present an Uninformative prior on *γ* over [0, ln 5].

**Figure S3:**
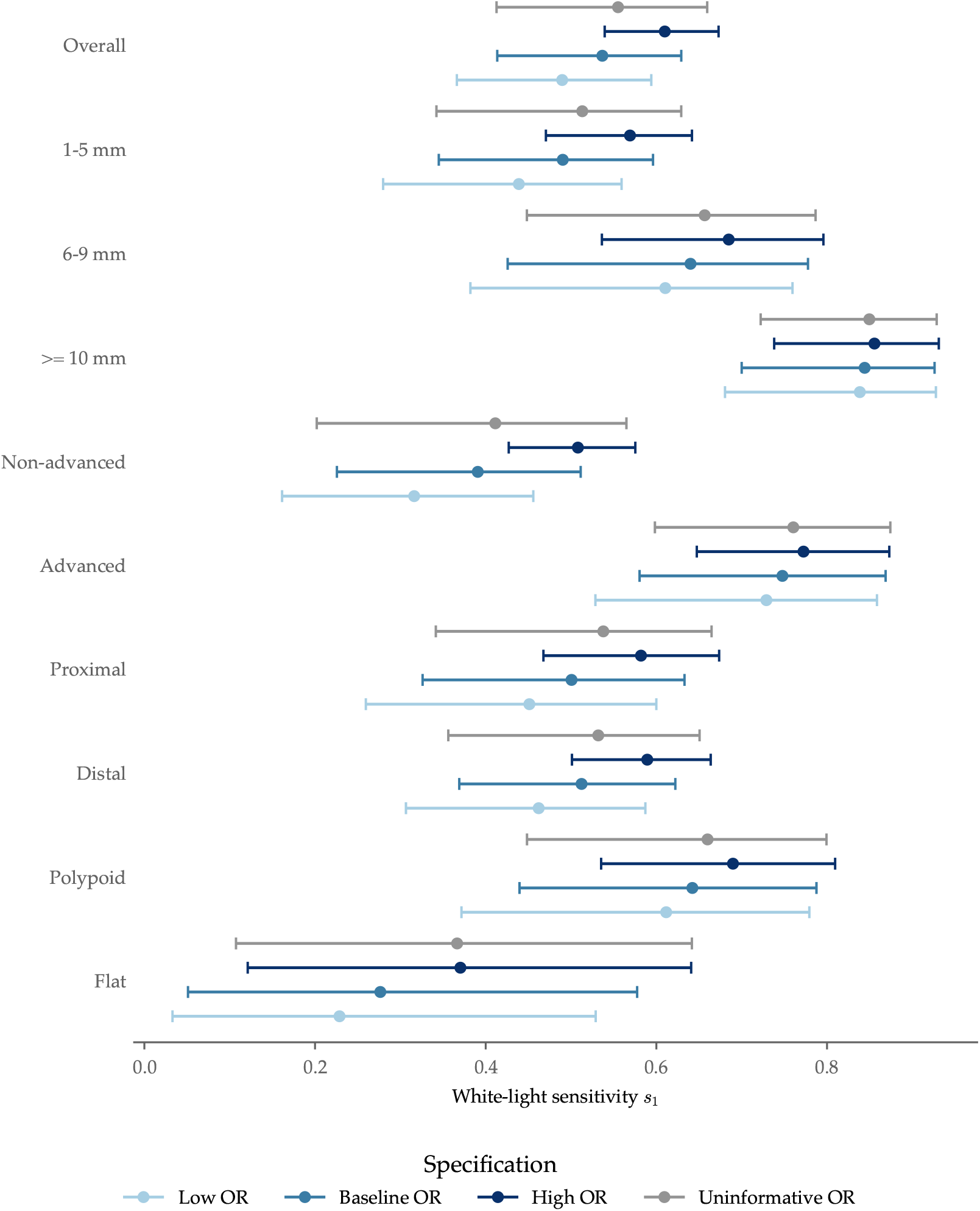
WLC sensitivity rises with the assumed detection odds ratio. *Notes:* WLC sensitivity *s*_1_ by subgroup. Posterior mean and 95% credible interval under each of the four prior specifications.

**Figure S4:**
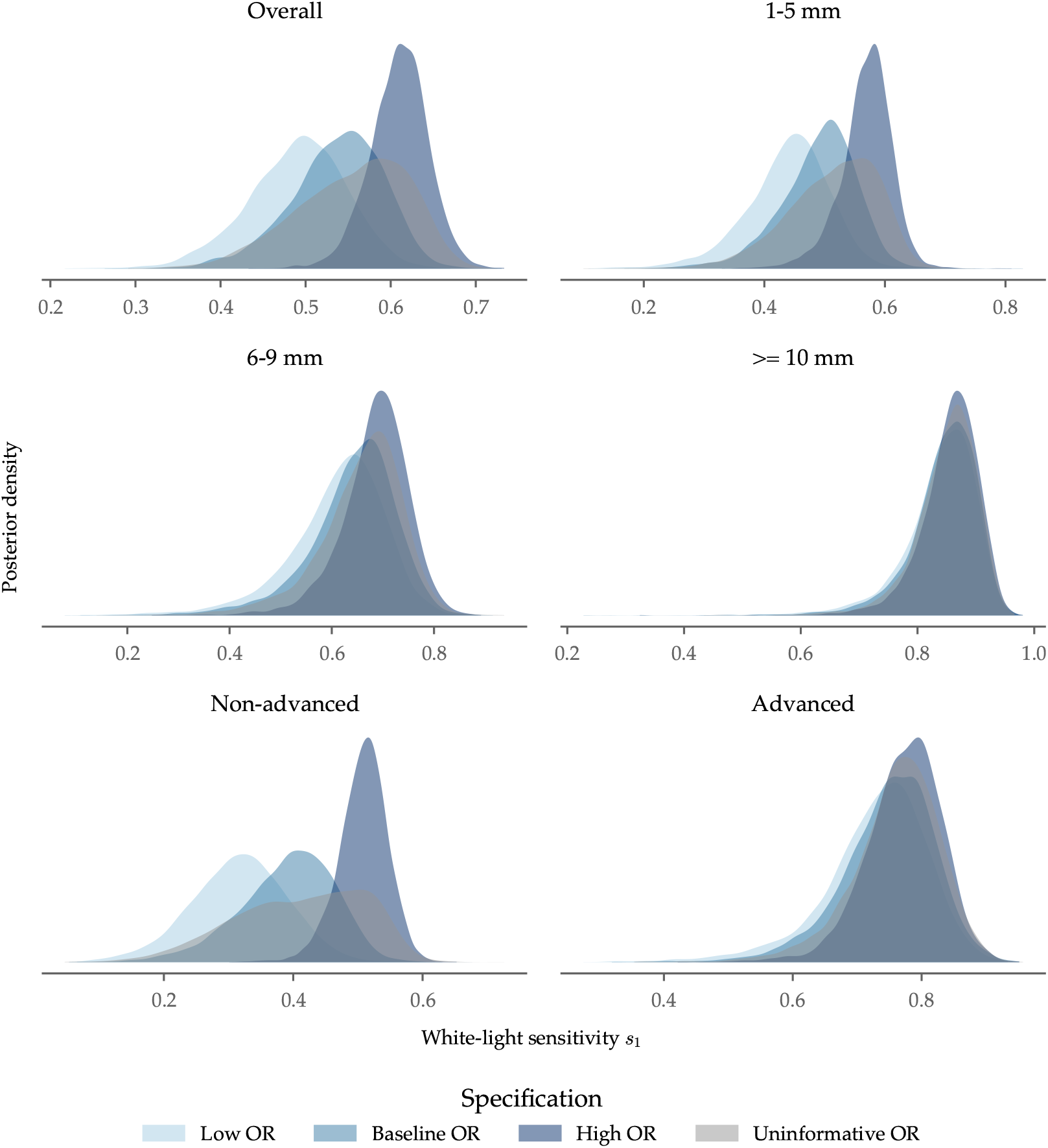
WLC sensitivity posteriors separate across specifications. *Notes:* WLC sensitivity posteriors under all four prior specifications, for six subgroups (overall, the three size bands, and the two histology groups.

**Figure S5:**
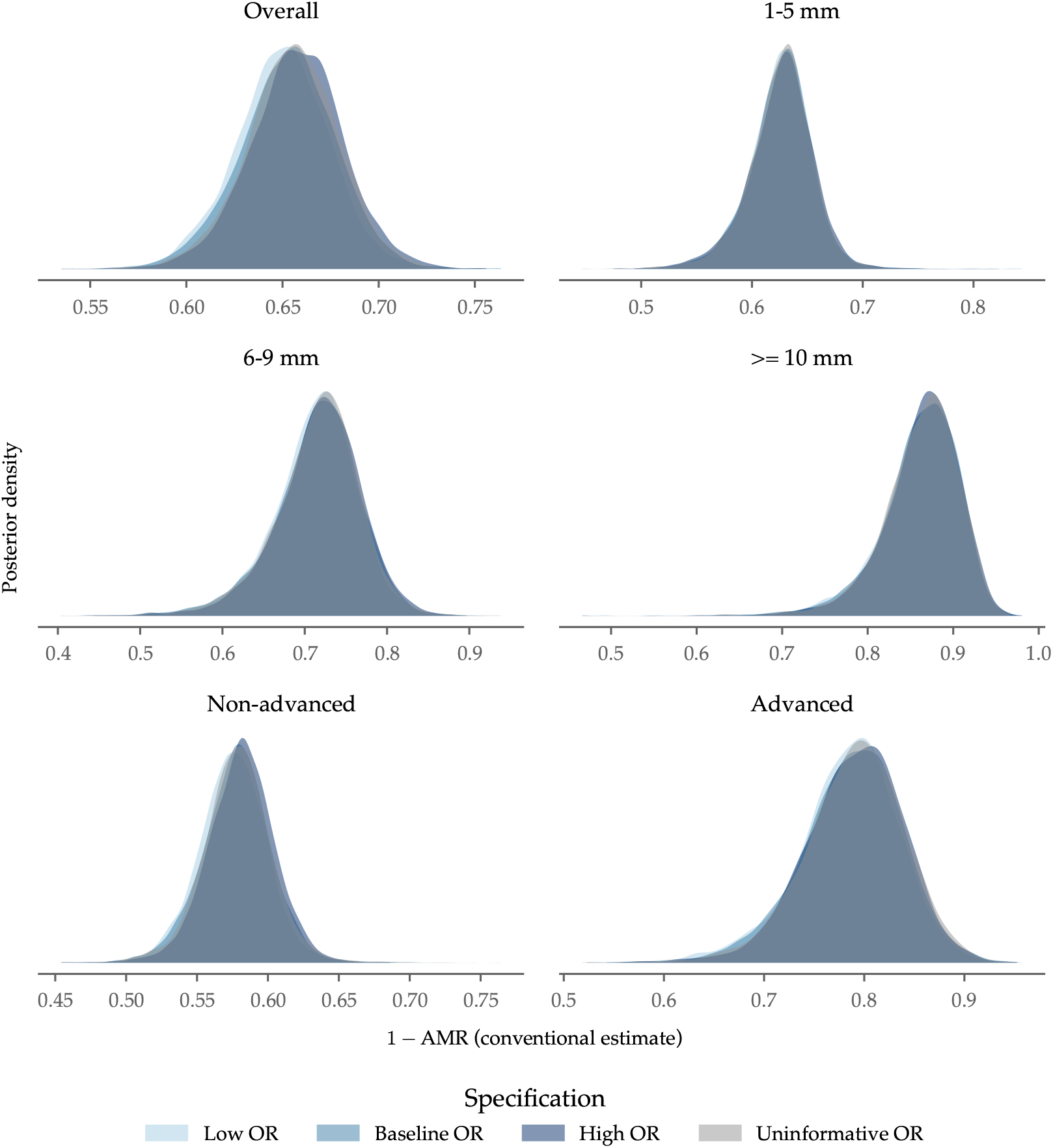
The model-implied miss rate is stable across specifications. *Notes:* Population 1 − AMR posteriors under all four prior distribution specifications for six of the ten subgroups.

**Table S1:**
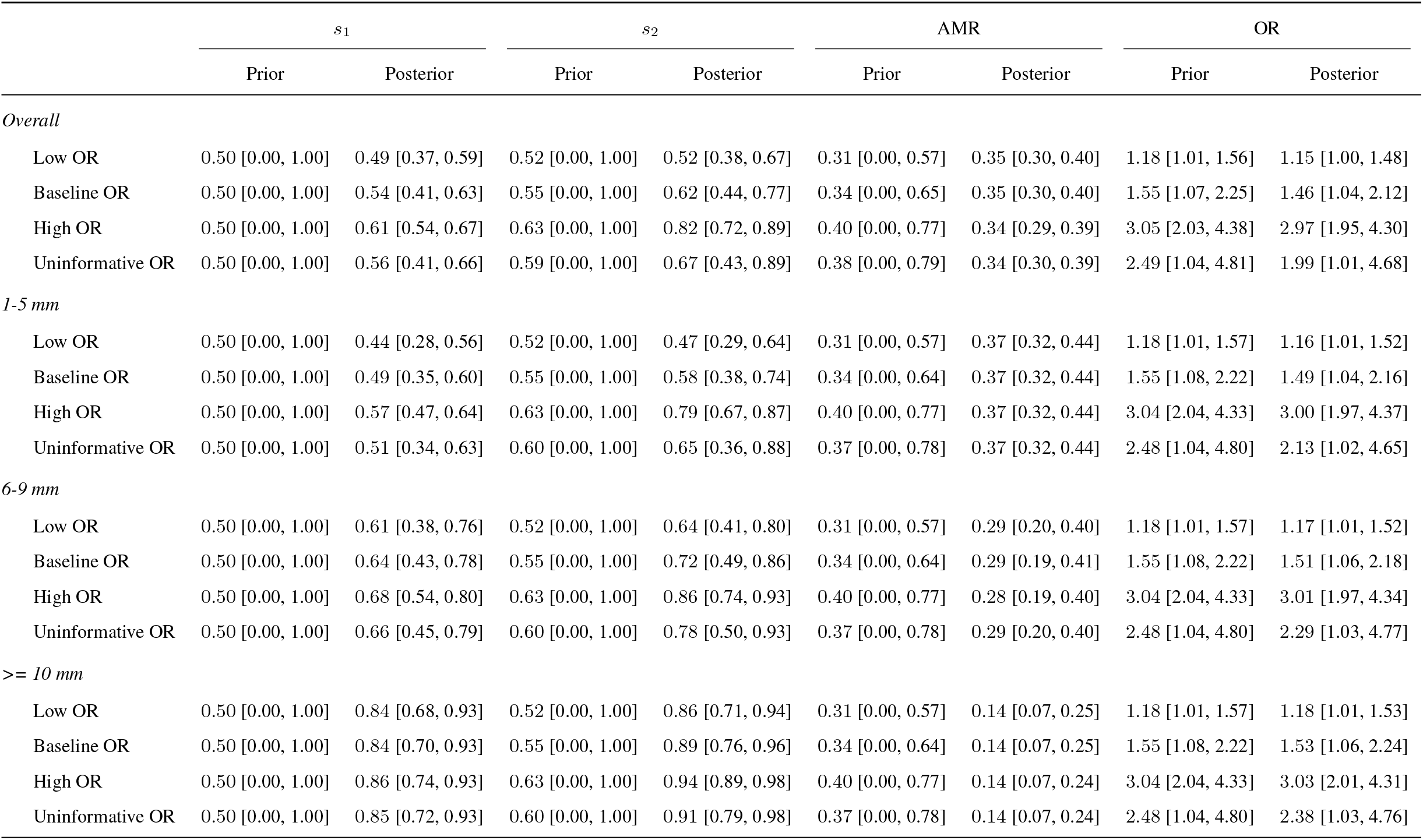
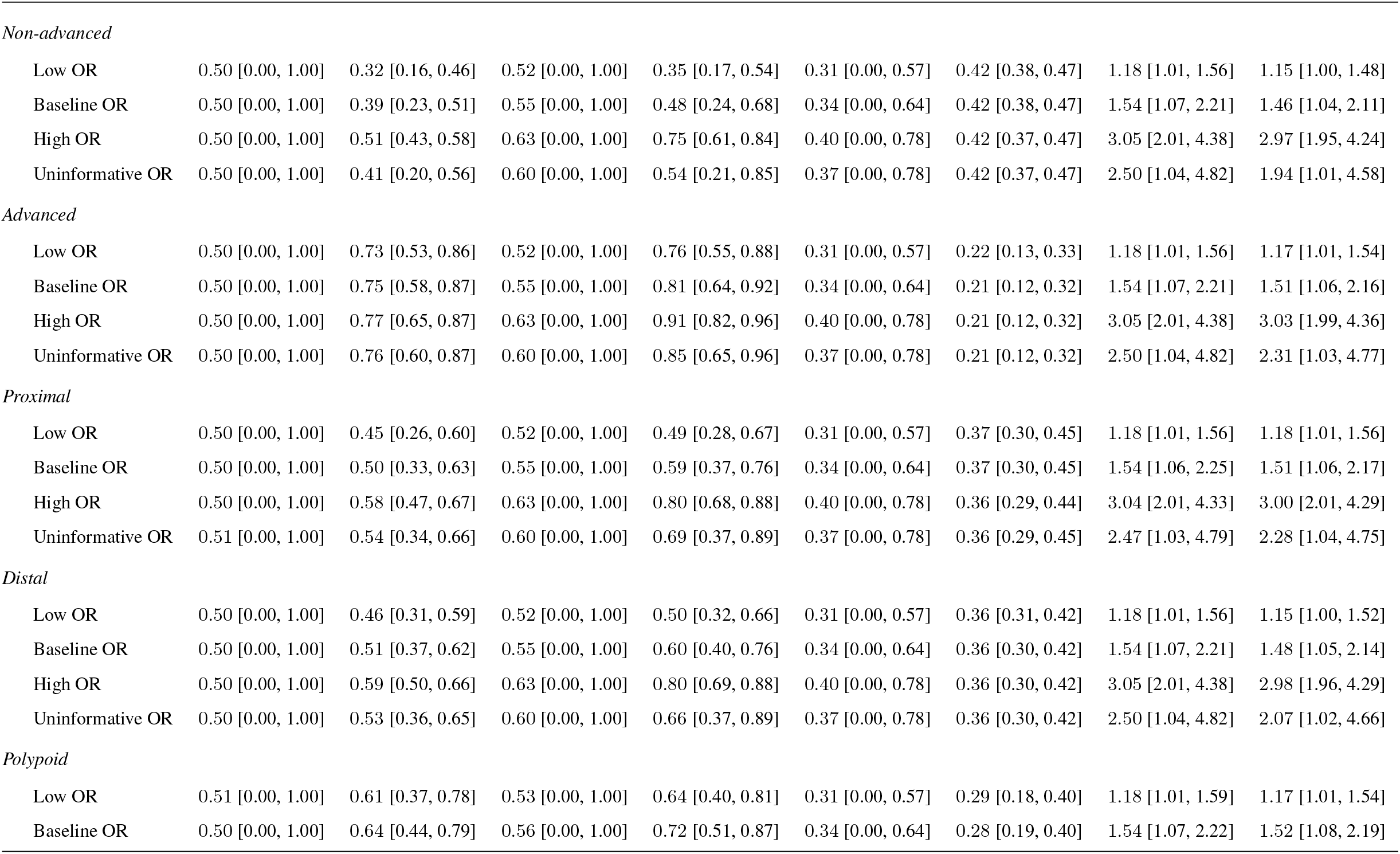
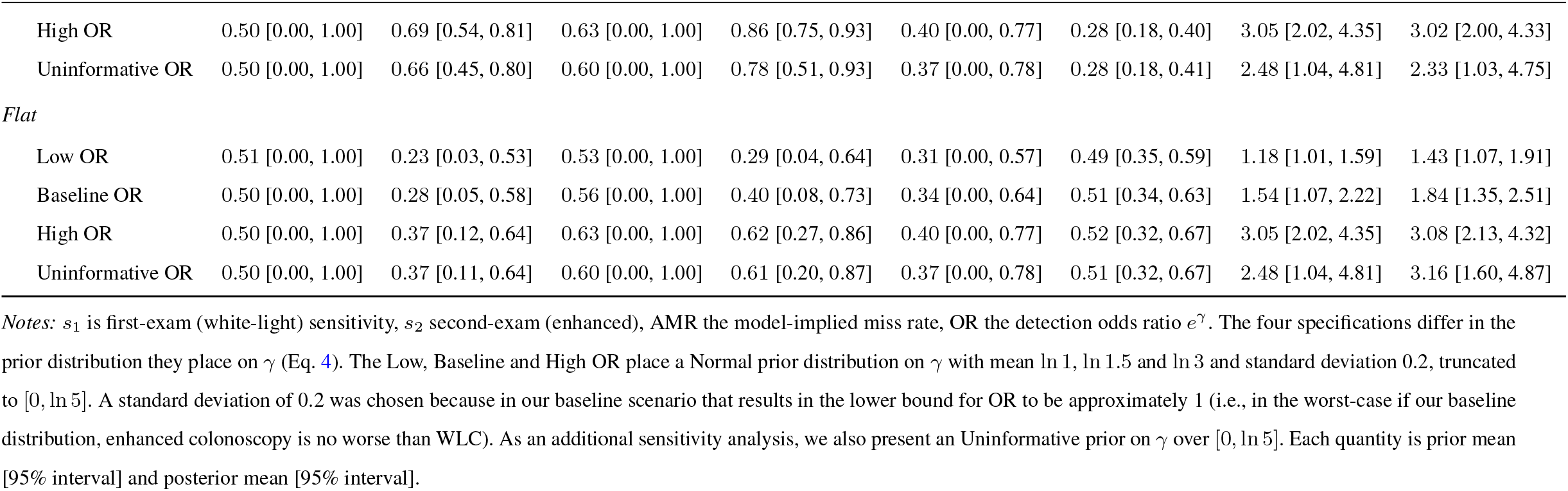
Population sensitivity by subgroup and specification, prior vs. posterior.

| | $s_1$ | | $s_2$ | | AMR | | OR | |
| --- | --- | --- | --- | --- | --- | --- | --- | --- |
|  | Prior | Posterior | Prior | Posterior | Prior | Posterior | Prior | Posterior |
| *Overall* |  |  |  |  |  |  |  |  |
| Low OR | 0.50 [0.00, 1.00] | 0.49 [0.37, 0.59] | 0.52 [0.00, 1.00] | 0.52 [0.38, 0.67] | 0.31 [0.00, 0.57] | 0.35 [0.30, 0.40] | 1.18 [1.01, 1.56] | 1.15 [1.00, 1.48] |
| Baseline OR | 0.50 [0.00, 1.00] | 0.54 [0.41, 0.63] | 0.55 [0.00, 1.00] | 0.62 [0.44, 0.77] | 0.34 [0.00, 0.65] | 0.35 [0.30, 0.40] | 1.55 [1.07, 2.25] | 1.46 [1.04, 2.12] |
| High OR | 0.50 [0.00, 1.00] | 0.61 [0.54, 0.67] | 0.63 [0.00, 1.00] | 0.82 [0.72, 0.89] | 0.40 [0.00, 0.77] | 0.34 [0.29, 0.39] | 3.05 [2.03, 4.38] | 2.97 [1.95, 4.30] |
| Uninformative OR | 0.50 [0.00, 1.00] | 0.56 [0.41, 0.66] | 0.59 [0.00, 1.00] | 0.67 [0.43, 0.89] | 0.38 [0.00, 0.79] | 0.34 [0.30, 0.39] | 2.49 [1.04, 4.81] | 1.99 [1.01, 4.68] |
| *1-5 mm* |  |  |  |  |  |  |  |  |
| Low OR | 0.50 [0.00, 1.00] | 0.44 [0.28, 0.56] | 0.52 [0.00, 1.00] | 0.47 [0.29, 0.64] | 0.31 [0.00, 0.57] | 0.37 [0.32, 0.44] | 1.18 [1.01, 1.57] | 1.16 [1.01, 1.52] |
| Baseline OR | 0.50 [0.00, 1.00] | 0.49 [0.35, 0.60] | 0.55 [0.00, 1.00] | 0.58 [0.38, 0.74] | 0.34 [0.00, 0.64] | 0.37 [0.32, 0.44] | 1.55 [1.08, 2.22] | 1.49 [1.04, 2.16] |
| High OR | 0.50 [0.00, 1.00] | 0.57 [0.47, 0.64] | 0.63 [0.00, 1.00] | 0.79 [0.67, 0.87] | 0.40 [0.00, 0.77] | 0.37 [0.32, 0.44] | 3.04 [2.04, 4.33] | 3.00 [1.97, 4.37] |
| Uninformative OR | 0.50 [0.00, 1.00] | 0.51 [0.34, 0.63] | 0.60 [0.00, 1.00] | 0.65 [0.36, 0.88] | 0.37 [0.00, 0.78] | 0.37 [0.32, 0.44] | 2.48 [1.04, 4.80] | 2.13 [1.02, 4.65] |
| *6-9 mm* |  |  |  |  |  |  |  |  |
| Low OR | 0.50 [0.00, 1.00] | 0.61 [0.38, 0.76] | 0.52 [0.00, 1.00] | 0.64 [0.41, 0.80] | 0.31 [0.00, 0.57] | 0.29 [0.20, 0.40] | 1.18 [1.01, 1.57] | 1.17 [1.01, 1.52] |
| Baseline OR | 0.50 [0.00, 1.00] | 0.64 [0.43, 0.78] | 0.55 [0.00, 1.00] | 0.72 [0.49, 0.86] | 0.34 [0.00, 0.64] | 0.29 [0.19, 0.41] | 1.55 [1.08, 2.22] | 1.51 [1.06, 2.18] |
| High OR | 0.50 [0.00, 1.00] | 0.68 [0.54, 0.80] | 0.63 [0.00, 1.00] | 0.86 [0.74, 0.93] | 0.40 [0.00, 0.77] | 0.28 [0.19, 0.40] | 3.04 [2.04, 4.33] | 3.01 [1.97, 4.34] |
| Uninformative OR | 0.50 [0.00, 1.00] | 0.66 [0.45, 0.79] | 0.60 [0.00, 1.00] | 0.78 [0.50, 0.93] | 0.37 [0.00, 0.78] | 0.29 [0.20, 0.40] | 2.48 [1.04, 4.80] | 2.29 [1.03, 4.77] |
| *>= 10 mm* |  |  |  |  |  |  |  |  |
| Low OR | 0.50 [0.00, 1.00] | 0.84 [0.68, 0.93] | 0.52 [0.00, 1.00] | 0.86 [0.71, 0.94] | 0.31 [0.00, 0.57] | 0.14 [0.07, 0.25] | 1.18 [1.01, 1.57] | 1.18 [1.01, 1.53] |
| Baseline OR | 0.50 [0.00, 1.00] | 0.84 [0.70, 0.93] | 0.55 [0.00, 1.00] | 0.89 [0.76, 0.96] | 0.34 [0.00, 0.64] | 0.14 [0.07, 0.25] | 1.55 [1.08, 2.22] | 1.53 [1.06, 2.24] |
| High OR | 0.50 [0.00, 1.00] | 0.86 [0.74, 0.93] | 0.63 [0.00, 1.00] | 0.94 [0.89, 0.98] | 0.40 [0.00, 0.77] | 0.14 [0.07, 0.24] | 3.04 [2.04, 4.33] | 3.03 [2.01, 4.31] |
| Uninformative OR | 0.50 [0.00, 1.00] | 0.85 [0.72, 0.93] | 0.60 [0.00, 1.00] | 0.91 [0.79, 0.98] | 0.37 [0.00, 0.78] | 0.14 [0.07, 0.24] | 2.48 [1.04, 4.80] | 2.38 [1.03, 4.76] |
|  | Prior | Posterior | Prior | Posterior | Prior | Posterior | Prior | Posterior |
| *Non-advanced* |  |  |  |  |  |  |  |  |
| Low OR | 0.50 [0.00, 1.00] | 0.32 [0.16, 0.46] | 0.52 [0.00, 1.00] | 0.35 [0.17, 0.54] | 0.31 [0.00, 0.57] | 0.42 [0.38, 0.47] | 1.18 [1.01, 1.56] | 1.15 [1.00, 1.48] |
| Baseline OR | 0.50 [0.00, 1.00] | 0.39 [0.23, 0.51] | 0.55 [0.00, 1.00] | 0.48 [0.24, 0.68] | 0.34 [0.00, 0.64] | 0.42 [0.38, 0.47] | 1.54 [1.07, 2.21] | 1.46 [1.04, 2.11] |
| High OR | 0.50 [0.00, 1.00] | 0.51 [0.43, 0.58] | 0.63 [0.00, 1.00] | 0.75 [0.61, 0.84] | 0.40 [0.00, 0.78] | 0.42 [0.37, 0.47] | 3.05 [2.01, 4.38] | 2.97 [1.95, 4.24] |
| Uninformative OR | 0.50 [0.00, 1.00] | 0.41 [0.20, 0.56] | 0.60 [0.00, 1.00] | 0.54 [0.21, 0.85] | 0.37 [0.00, 0.78] | 0.42 [0.37, 0.47] | 2.50 [1.04, 4.82] | 1.94 [1.01, 4.58] |
| *Advanced* |  |  |  |  |  |  |  |  |
| Low OR | 0.50 [0.00, 1.00] | 0.73 [0.53, 0.86] | 0.52 [0.00, 1.00] | 0.76 [0.55, 0.88] | 0.31 [0.00, 0.57] | 0.22 [0.13, 0.33] | 1.18 [1.01, 1.56] | 1.17 [1.01, 1.54] |
| Baseline OR | 0.50 [0.00, 1.00] | 0.75 [0.58, 0.87] | 0.55 [0.00, 1.00] | 0.81 [0.64, 0.92] | 0.34 [0.00, 0.64] | 0.21 [0.12, 0.32] | 1.54 [1.07, 2.21] | 1.51 [1.06, 2.16] |
| High OR | 0.50 [0.00, 1.00] | 0.77 [0.65, 0.87] | 0.63 [0.00, 1.00] | 0.91 [0.82, 0.96] | 0.40 [0.00, 0.78] | 0.21 [0.12, 0.32] | 3.05 [2.01, 4.38] | 3.03 [1.99, 4.36] |
| Uninformative OR | 0.50 [0.00, 1.00] | 0.76 [0.60, 0.87] | 0.60 [0.00, 1.00] | 0.85 [0.65, 0.96] | 0.37 [0.00, 0.78] | 0.21 [0.12, 0.32] | 2.50 [1.04, 4.82] | 2.31 [1.03, 4.77] |
| *Proximal* |  |  |  |  |  |  |  |  |
| Low OR | 0.50 [0.00, 1.00] | 0.45 [0.26, 0.60] | 0.52 [0.00, 1.00] | 0.49 [0.28, 0.67] | 0.31 [0.00, 0.57] | 0.37 [0.30, 0.45] | 1.18 [1.01, 1.56] | 1.18 [1.01, 1.56] |
| Baseline OR | 0.50 [0.00, 1.00] | 0.50 [0.33, 0.63] | 0.55 [0.00, 1.00] | 0.59 [0.37, 0.76] | 0.34 [0.00, 0.64] | 0.37 [0.30, 0.45] | 1.54 [1.06, 2.25] | 1.51 [1.06, 2.17] |
| High OR | 0.50 [0.00, 1.00] | 0.58 [0.47, 0.67] | 0.63 [0.00, 1.00] | 0.80 [0.68, 0.88] | 0.40 [0.00, 0.78] | 0.36 [0.29, 0.44] | 3.04 [2.01, 4.33] | 3.00 [2.01, 4.29] |
| Uninformative OR | 0.51 [0.00, 1.00] | 0.54 [0.34, 0.66] | 0.60 [0.00, 1.00] | 0.69 [0.37, 0.89] | 0.37 [0.00, 0.78] | 0.36 [0.29, 0.45] | 2.47 [1.03, 4.79] | 2.28 [1.04, 4.75] |
| *Distal* |  |  |  |  |  |  |  |  |
| Low OR | 0.50 [0.00, 1.00] | 0.46 [0.31, 0.59] | 0.52 [0.00, 1.00] | 0.50 [0.32, 0.66] | 0.31 [0.00, 0.57] | 0.36 [0.31, 0.42] | 1.18 [1.01, 1.56] | 1.15 [1.00, 1.52] |
| Baseline OR | 0.50 [0.00, 1.00] | 0.51 [0.37, 0.62] | 0.55 [0.00, 1.00] | 0.60 [0.40, 0.76] | 0.34 [0.00, 0.64] | 0.36 [0.30, 0.42] | 1.54 [1.07, 2.21] | 1.48 [1.05, 2.14] |
| High OR | 0.50 [0.00, 1.00] | 0.59 [0.50, 0.66] | 0.63 [0.00, 1.00] | 0.80 [0.69, 0.88] | 0.40 [0.00, 0.78] | 0.36 [0.30, 0.42] | 3.05 [2.01, 4.38] | 2.98 [1.96, 4.29] |
| Uninformative OR | 0.50 [0.00, 1.00] | 0.53 [0.36, 0.65] | 0.60 [0.00, 1.00] | 0.66 [0.37, 0.89] | 0.37 [0.00, 0.78] | 0.36 [0.30, 0.42] | 2.50 [1.04, 4.82] | 2.07 [1.02, 4.66] |
| *Polypoid* |  |  |  |  |  |  |  |  |
| Low OR | 0.51 [0.00, 1.00] | 0.61 [0.37, 0.78] | 0.53 [0.00, 1.00] | 0.64 [0.40, 0.81] | 0.31 [0.00, 0.57] | 0.29 [0.18, 0.40] | 1.18 [1.01, 1.59] | 1.17 [1.01, 1.54] |
| Baseline OR | 0.50 [0.00, 1.00] | 0.64 [0.44, 0.79] | 0.56 [0.00, 1.00] | 0.72 [0.51, 0.87] | 0.34 [0.00, 0.64] | 0.28 [0.19, 0.40] | 1.54 [1.07, 2.22] | 1.52 [1.08, 2.19] |
|  | Prior | Posterior | Prior | Posterior | Prior | Posterior | Prior | Posterior |
| High OR | 0.50 [0.00, 1.00] | 0.69 [0.54, 0.81] | 0.63 [0.00, 1.00] | 0.86 [0.75, 0.93] | 0.40 [0.00, 0.77] | 0.28 [0.18, 0.40] | 3.05 [2.02, 4.35] | 3.02 [2.00, 4.33] |
| Uninformative OR | 0.50 [0.00, 1.00] | 0.66 [0.45, 0.80] | 0.60 [0.00, 1.00] | 0.78 [0.51, 0.93] | 0.37 [0.00, 0.78] | 0.28 [0.18, 0.41] | 2.48 [1.04, 4.81] | 2.33 [1.03, 4.75] |
| *Flat* |  |  |  |  |  |  |  |  |
| Low OR | 0.51 [0.00, 1.00] | 0.23 [0.03, 0.53] | 0.53 [0.00, 1.00] | 0.29 [0.04, 0.64] | 0.31 [0.00, 0.57] | 0.49 [0.35, 0.59] | 1.18 [1.01, 1.59] | 1.43 [1.07, 1.91] |
| Baseline OR | 0.50 [0.00, 1.00] | 0.28 [0.05, 0.58] | 0.56 [0.00, 1.00] | 0.40 [0.08, 0.73] | 0.34 [0.00, 0.64] | 0.51 [0.34, 0.63] | 1.54 [1.07, 2.22] | 1.84 [1.35, 2.51] |
| High OR | 0.50 [0.00, 1.00] | 0.37 [0.12, 0.64] | 0.63 [0.00, 1.00] | 0.62 [0.27, 0.86] | 0.40 [0.00, 0.77] | 0.52 [0.32, 0.67] | 3.05 [2.02, 4.35] | 3.08 [2.13, 4.32] |
| Uninformative OR | 0.50 [0.00, 1.00] | 0.37 [0.11, 0.64] | 0.60 [0.00, 1.00] | 0.61 [0.20, 0.87] | 0.37 [0.00, 0.78] | 0.51 [0.32, 0.67] | 2.48 [1.04, 4.81] | 3.16 [1.60, 4.87] |

Notes: $s_1$ is first-exam (white-light) sensitivity, $s_2$ second-exam (enhanced), AMR the model-implied miss rate, OR the detection odds ratio $e^\gamma$ . The four specifications differ in the prior distribution they place on $\gamma$ (Eq. 4). The Low, Baseline and High OR place a Normal prior distribution on $\gamma$ with mean $\ln 1$ , $\ln 1.5$ and $\ln 3$ and standard deviation 0.2, truncated to $[0, \ln 5]$ . A standard deviation of 0.2 was chosen because in our baseline scenario that results in the lower bound for OR to be approximately 1 (i.e., in the worst-case if our baseline distribution, enhanced colonoscopy is no worse than WLC). As an additional sensitivity analysis, we also present an Uninformative prior on $\gamma$ over $[0, \ln 5]$ . Each quantity is prior mean [95% interval] and posterior mean [95% interval].

## References and Notes

[1] Amy B. Knudsen, Carolyn M. Rutter, Elisabeth F.P. Peterse, Anna P. Lietz, Claudia L. Seguin, Reinier G.S. Meester, Leslie A. Perdue, Jennifer S. Lin, Rebecca L. Siegel, V. Paul Doria-Rose, Eric J. Feuer, Ann G. Zauber, Karen M. Kuntz, and Iris Lansdorp-Vogelaar. Colorectal Cancer Screening: An Updated Modeling Study for the US Preventive Services Task Force. JAMA - Journal of the American Medical Association, 325(19):1998–2011, 2021. doi: 10.1001/jama.2021.5746.

[2] Martin C. S. Wong, Junjie Huang, Jason L. W. Huang, Tiffany W. Y. Pang, Peter Choi, Jingxuan Wang, Jason I. Chiang, and Johnny Yu Jiang. Global Prevalence of Colorectal Neoplasia: A Systematic Review and Meta-Analysis. Clinical Gastroenterology and Hepatology, 18(3):553–561.e10, March 2020. ISSN 1542-3565, 1542-7714. doi: 10.1016/j.cgh.2019.07.016.

[3] Pedro Nascimento de Lima, Christopher Maerzluft, Jonathan Ozik, Nicholson Collier, and Carolyn M. Rutter. Stress-Testing US Colorectal Cancer Screening Guidelines: Decennial Colonoscopy from Age 45 is Robust to Natural History Uncertainty and Colonoscopy Sensitivity Assumptions. Medical Decision Making, 45(5):557–568, July 2025. ISSN 0272-989X. doi: 10.1177/0272989X251334373.

[4] Jeroen C. Van Rijn, Johannes B. Reitsma, Jaap Stoker, Patrick M. Bossuyt, Sander J. Van Deventer, and Evelien Dekker. Polyp miss rate determined by tandem colonoscopy: A systematic review. American Journal of Gastroenterology, 101(2):343–350, 2006. doi: 10.1111/j.1572-0241.2006.00390.x.

[5] Shengbing Zhao, Shuling Wang, Peng Pan, Tian Xia, Xin Chang, Xia Yang, Liliangzi Guo, Qianqian Meng, Fan Yang, Wei Qian, Zhichao Xu, Yuanqiong Wang, Zhijie Wang, Lun Gu, Rundong Wang, Fangzhou Jia, Jun Yao, Zhaoshen Li, and Yu Bai. Magnitude, Risk Factors, and Factors Associated With Adenoma Miss Rate of Tandem Colonoscopy: A Systematic Review and Meta-analysis. Gastroenterology, 156(6):1661–1674.e11, 2019. doi: 10.1053/j.gastro.2019.01.260.

[6] Beate Jahn, Marvin Bundo, Marjan Arvandi, Monika Schaffner, Jovan Todorovic, Gaby Sroczynski, Amy Knudsen, Timo Fischer, Irmgard Schiller-Fruehwirth, Dietmar Öfner, Friedrich Renner, Michael Jonas, Igor Kuchin, Julia Kruse, Júlia Santamaria, Monika Ferlitsch, and Uwe Siebert. One in three adenomas could be missed by white-light colonoscopy – findings from a systematic review and meta-analysis. BMC Gastroenterology, 25(1):170, March 2025. ISSN 1471-230X. doi: 10.1186/s12876-025-03679-4.

[7] Elie Tamer. Partial Identification in Econometrics. Annual Review of Economics, 2(1):167–195, September 2010. ISSN 1941-1383, 1941-1391. doi: 10.1146/annurev.economics.050708.143401.

[8] Raffaella Giacomini and Toru Kitagawa. Robust Bayesian Inference for Set-Identified Models. Econometrica, 89 (4):1519–1556, 2021. ISSN 0012-9682. doi: 10.3982/ECTA16773.

[9] Omiros Papaspiliopoulos, Gareth O. Roberts, and Martin Sköld. A General Framework for the Parametrization of Hierarchical Models. Statistical Science, 22(1):59–73, February 2007. ISSN 0883-4237, 2168-8745. doi: 10.1214/088342307000000014.

[10] Michael Betancourt and Mark Girolami. Hamiltonian Monte Carlo for Hierarchical Models. In Current Trends in Bayesian Methodology with Applications. Chapman and Hall/CRC, 2015. doi: 10.1201/b18502-5.

[11] Daniel Castaneda, Violeta B. Popov, Elijah Verheyen, Praneet Wander, and Seth A. Gross. New technologies improve adenoma detection rate, adenoma miss rate, and polyp detection rate: A systematic review and meta-analysis. Gastrointestinal Endoscopy, 88(2):209–222.e11, August 2018. ISSN 0016-5107. doi: 10.1016/j.gie.2018.03.022.

[12] Bob Carpenter, Andrew Gelman, Matthew D. Hoffman, Daniel Lee, Ben Goodrich, Michael Betancourt, Marcus Brubaker, Jiqiang Guo, Peter Li, and Allen Riddell. Stan : A Probabilistic Programming Language. Journal of Statistical Software, 76(1):1–32, 2017. ISSN 1548-7660. doi: 10.18637/jss.v076.i01.

[13] Matthew D. Hoffman and Andrew Gelman. The No-U-Turn Sampler: Adaptively Setting Path Lengths in Hamiltonian Monte Carlo. Journal of Machine Learning Research, 15(47):1593–1623, 2014. ISSN 1533-7928.

[14] Aki Vehtari, Andrew Gelman, Daniel Simpson, Bob Carpenter, and Paul-Christian Bürkner. Rank-Normalization, Folding, and Localization: An Improved R^ for Assessing Convergence of MCMC (with Discussion). Bayesian Analysis, 16(2):667–718, June 2021. ISSN 1936-0975. doi: 10.1214/20-BA1221.

[15] Ann Zauber, Iris Lansdorp-Vogelaar, Amy B Knudsen, Janneke Wilschut, Marjolein van Ballegooijen, and Kun. Annals of Internal Medicine Clinical Guidelines Evaluating Test Strategies for Colorectal Cancer Screening :. Annals of internal medicine, 149(9):659–669, 2008.

[16] Amy B. Knudsen, Ann G. Zauber, Carolyn M. Rutter, Steffie K. Naber, V. Paul Doria-Rose, Chester Pabiniak, Colden Johanson, Sara E. Fischer, Iris Lansdorp-Vogelaar, and Karen M. Kuntz. Estimation of benefits, burden, and harms of colorectal cancer screening strategies: Modeling study for the US preventive services Task Force. JAMA - Journal of the American Medical Association, 315(23):2595–2609, 2016. doi: 10.1001/jama.2016.6828.

[17] Cesare Hassan, Marco Spadaccini, Yuichi Mori, Farid Foroutan, Antonio Facciorusso, Paraskevas Gkolfakis, Georgios Tziatzios, Konstantinos Triantafyllou, Giulio Antonelli, Kareem Khalaf, Tommy Rizkala, Per Olav Vandvik, Alessandro Fugazza, Emanuele Rondonotti, Jeremy R. Glissen-Brown, Shunsuke Kamba, Marcello Maida, Loredana Correale, Pradeep Bhandari, Rodrigo Jover, Prateek Sharma, Douglas K. Rex, and Alessandro Repici. Real-Time Computer-Aided Detection of Colorectal Neoplasia During Colonoscopy: A Systematic Review and Meta-analysis. Annals of Internal Medicine, 176(9):1209–1220, September 2023. ISSN 0003-4819, 1539-3704. doi: 10.7326/M22-3678.

[18] Brian T Clark, Tarun Rustagi, and Loren Laine. What Level of Bowel Prep Quality Requires Early Repeat Colonoscopy: Systematic Review and Meta-Analysis of the Impact of Preparation Quality on Adenoma Detection Rate. American Journal of Gastroenterology, 109(11):1714–1723, November 2014. ISSN 0002-9270. doi: 10.1038/ajg.2014.232.

[19] Kevin O. Turner, Robert M. Genta, and Amnon Sonnenberg. Lesions of all types exist in colon polyps of all sizes. American Journal of Gastroenterology, 113(2):303–306, 2018. doi: 10.1038/ajg.2017.439.

[20] H. Brenner, C. Stock, and M. Hoffmeister. Effect of screening sigmoidoscopy and screening colonoscopy on colorectal cancer incidence and mortality: Systematic review and meta-analysis of randomised controlled trials and observational studies. BMJ, 348(apr09 1):g2467–g2467, April 2014. ISSN 1756-1833. doi: 10.1136/bmj.g2467.

[21] Pedro Nascimento de Lima, Laura Matrajt, Gloria Coronado, Anne L. Escaron, and Carolyn M. Rutter. Cost-Effectiveness of Noninvasive Colorectal Cancer Screening in Community Clinics. JAMA Network Open, 8(1): e2454938, January 2025. ISSN 2574-3805. doi: 10.1001/jamanetworkopen.2024.54938.

[22] Lalitha Sanathanan. Estimating the Size of a Multinomial Population. The Annals of Mathematical Statistics, 43 (1):142–152, February 1972. ISSN 0003-4851, 2168-8990. doi: 10.1214/aoms/1177692709.

